# Longitudinal associations of behavioral problems and text reading skills in school-aged children born term and preterm

**DOI:** 10.1101/2021.10.30.21265689

**Authors:** Machiko Hosoki, Lauren Borchers, Virginia A. Marchman, Katherine E. Travis, Heidi M. Feldman

## Abstract

We assessed the contribution of total behavioral problems at 6 years to text reading skills at 8 years in children born term and preterm. Birth group moderated associations among total behavioral problems and reading skills; total behavioral problems predicted reading skills in the term but not preterm group.

## Introduction

Reading skills vary widely in school-aged children. Late reading abilities are associated with functioning in several other domains in younger age, including language, phonological awareness, executive function, and non-verbal intelligence[1]. Performance in associated domains in early development has been found to improve prediction of individual differences in later reading skills[2].

Children born preterm are at risk for long-term developmental decrements in reading skills[3]. We previously reported that language and phonological awareness at age 6 predicted text reading at age 8 years in a sample of children born term and preterm[1]. However, executive function and non-verbal intelligence improved the prediction of text reading abilities at age 8 only in the preterm group[1]. These findings suggested that skills contributing to text reading abilities differed in the two birth groups.

In the current study, we expanded on findings reported by Borchers et al.[1] by considering the longitudinal prediction of behavioral problems to text reading skills in school-aged children born term and preterm. We quantified the severity of behavioral problems as the score on a parent-report measure that included internalizing problems (such as anxiety and depression), and externalizing problems (such as oppositionality and aggression)[4]. We assessed the strength of association of the behavioral problems score and measures of executive function and non-verbal intelligence to assure that we were tapping into a different construct underlying reading abilities than what was previously analyzed[1]. Based on the broader set of associations observed in children born preterm in the earlier study, we hypothesized that behavioral problems at age 6 would be negatively associated with text reading skills at age 8 in children born preterm, but not those born at term. We further anticipated that behavioral problems would uniquely contribute to later reading skills, and that the negative association would persist after consideration of individual differences in early reading skills.

## Materials and Methods

### Participants

Participants were enrolled in a longitudinal study of reading skills and white matter characteristics in children born at term and preterm. The characteristics of children enrolled have been described in the previous study[1]. Children were recruited from the San Francisco Bay Area from 2012 to 2015 and were followed from age 6 to 8 years. Preterm birth was restricted to children born at gestational age ≤ 32 weeks; these very and extremely preterm children are at high risk for problems in multiple domains of function, including behavioral problems and reading decrements[3]. These deficits have been attributed to a preterm encephalopathy that includes injury and dysmaturity of white matter pathways[5]. Full term birth was defined as gestational age ≥37 weeks or birth weight ≥2500 grams[1,3].

The experimental protocol was approved by the Stanford University Institutional Review Board. Written consent was obtained from a parent or a legal guardian. We collected demographic characteristics, including sex, birth group, and birthweight.

### Behavioral assessments

When the participants were age 6, parents completed the Child Behavior Checklist (CBCL)/6-18, a questionnaire that evaluates children’s behaviors[4]. The main predictor variable was the CBCL total problems standardized score, which includes internalizing problems, externalizing problems, thought problems, social problems and attention problems[4]. In the CBCL, behavioral problems were classified as clinically elevated if the standardized scores were at or above 60. Participants completed the Test of Word Reading Efficiency (TOWRE) 2^nd^ edition at age 6. The TOWRE-2 evaluates children’s decoding efficiency by asking the child to quickly read words and pseudowords. We controlled for early reading skills using scores on the TOWRE-2 reading efficiency test at age 6. Participants took the Gray Oral Reading Test-5 (GORT) at age 8, a standardized test that evaluates reading fluency and comprehension of text passages[6]. We used the Oral Reading Index as the primary outcome measure for text reading skills.

Participants performed three executive function tasks—Spatial Span, Spatial Working Memory, and Stockings of Cambridge—from the Cambridge Neuropsychological Test Automated Battery (CANTAB)[1]. We used a single factor from factor analysis to summarize their performance on the three tasks. The detailed description of computing this factor is described in Borchers et al[1]. We assessed non-verbal Intelligence Quotient (IQ) at age 6 using the Perceptual Reasoning Index from the Wechsler Abbreviated Scale of Intelligence-II (WASI-II)[1,7].

### Statistical Analysis

All statistical analyses were conducted using *R* with statistical significance set at *p*<0.05. For this analysis, we included children whose parents completed the CBCL, and who completed the TOWRE, the CANTAB, and the WASI-II at age 6, and the GORT at age 8. This sample is a subset of those children included in Borchers et al[1]. We conducted chi-squared tests to compare the proportion of sex, and the proportion of participants with clinically elevated CBCL total problems within children born at term and children born preterm. Using independent samples t-test, we compared CBCL total problems, TOWRE word reading efficiency, and GORT oral reading index by birth group. The Welch two sample t-test was utilized, when there were unequal variances. We assessed Pearson correlations of the CBCL total problems score with the executive function factor and with non-verbal IQ to assure that the measures were tapping distinct abilities.

We ran a series of linear regression models to assess the longitudinal contribution of behavioral problems to text reading skills and whether birth group played a moderating role. We assessed the contribution of sex, birth group, and behavioral problems at age 6 to text reading skills at age 8 (model 1A). We then added an interaction term of behavioral problems at age 6 and birth group (model 1B). Finally, we added early reading skills at age 6 to model 1B to evaluate whether the association between the behavioral problems at age 6 and text reading skills at age 8 differs by birth group after consideration of early reading skills (model 1C).

## Results

A total of 37 children born preterm (PT) (23 boys) and 40 children born at term (FT) (16 boys) were included in this study. Of 77 children, 15 children (PT: 9 children, FT: 6 children)’s CBCL total problems were clinical elevated. Sex proportion by birth group approached statistical significance (*X*^2^(1)=2.94, *p*=.086) and therefore sex was included in the regression models. The difference in proportions of children with elevated total problems in the two groups was not statistically significant (*X*^2^(1)=0.55, *p=*.457). By design, the mean birthweight was lower in children born PT than FT (PT: 1398.1±439.7 g, FT: 3298.5±408.6 g, t(75)=19.66, *p*<.001).

There were no birth group differences in CBCL total problems (PT: 50.6± 3.1, FT: 47.2 ±10.6, t(75)=-1.26, *p*=.212), TOWRE word reading efficiency (PT: 95.6±20.7, FT: 100.6±24.3, t(75)=0.964, *p*=.338), or GORT oral reading index (PT: 97.6±10.5, FT: 100.4 ±14.6, t(70.79)=0.93, *p*=.355).

Correlations between CBCL total problems and executive function at age 6 (r=-0.29, *p*=.012), and between CBCL total problems and non-verbal IQ (r=-0.35, *p=*.002) were weak. These analyses confirmed that measures from these domains were not tapping into the same underlying abilities that were previously assessed[1].

Table 1 demonstrates the results of linear regression models that investigated the longitudinal contributions of behavioral problems at age 6 to text reading skills at age 8. Model 1A demonstrates that sex, birth group, and behavioral problems at age 6 jointly contributed over 11% to text reading skills at age 8. Sex was the only unique predictor. Model 1B shows a significant increase in variance accounted for when the interaction term with birth group was added, reaching over 17%. Thus, the relation between behavioral problems and reading was significantly moderated by birth group. Model 1C shows that the addition of early reading skills further improved the model, accounting for over 50% of variance; birth group continued to moderate the association between behavioral problems at age 6 and text reading skills at age 8. Figure 1 illustrates the interaction of birth group and behavioral problems after early reading skills at age 6 were considered; the slope was significantly negative for the FT group and non-significant for the PT group.

**Table 1:** Predicting text reading skills at age 8 from behavioral total problems at age 6 and birth group controlling for reading skills at age 6

|  | Model 1A | Model 1B | Model 1C |
| --- | --- | --- | --- |
| Sex | 6.35 (2.87)* | 6.15 (2.79)* | 2.79 (2.21) |
| Birth Group | -0.54 (2.89) | -26.9 (11.9)* | -18.47 (9.27) |
| Behavioral Problems at 6<sup>a</sup> | -0.21 (0.12)<sup>#</sup> | -0.53 (0.18)** | -0.33 (0.14)* |
| Behavioral Problems at 6<sup>a</sup> x<br>Birth group |  | 0.54 (0.24)* | 0.38 (0.18)* |
| Early Text Reading Skills at 6<sup>b</sup> |  |  | **0.34 (0.05)\*\*** |
| $\Delta r^2$ | - | **6.0 % \*** | **33.6 %\*\*** |
| Total R<sup>2</sup> | **11.3%\*** | **17.3 %\*\*** | **50.9 %\*\*** |
| Adjusted R<sup>2</sup> | 7.6 %* | 12.7 %** | 47.5 %** |

Note: Data are non-standardized coefficients (SE). \* $p < 0.05$ \*\* $p < 0.01$ , # $p < 0.10$ . The $r^2$ changes for models 1B and C are in reference to the preceding model. <sup>a</sup> Behavioral problems at 6, measured with the CBCL total problems standardized score. <sup>b</sup> Early reading skills at 6, measured with the TOWRE reading efficiency standardized score. Text reading skills at age 8, measured with the GORT Oral reading index.

**Figure 1.**
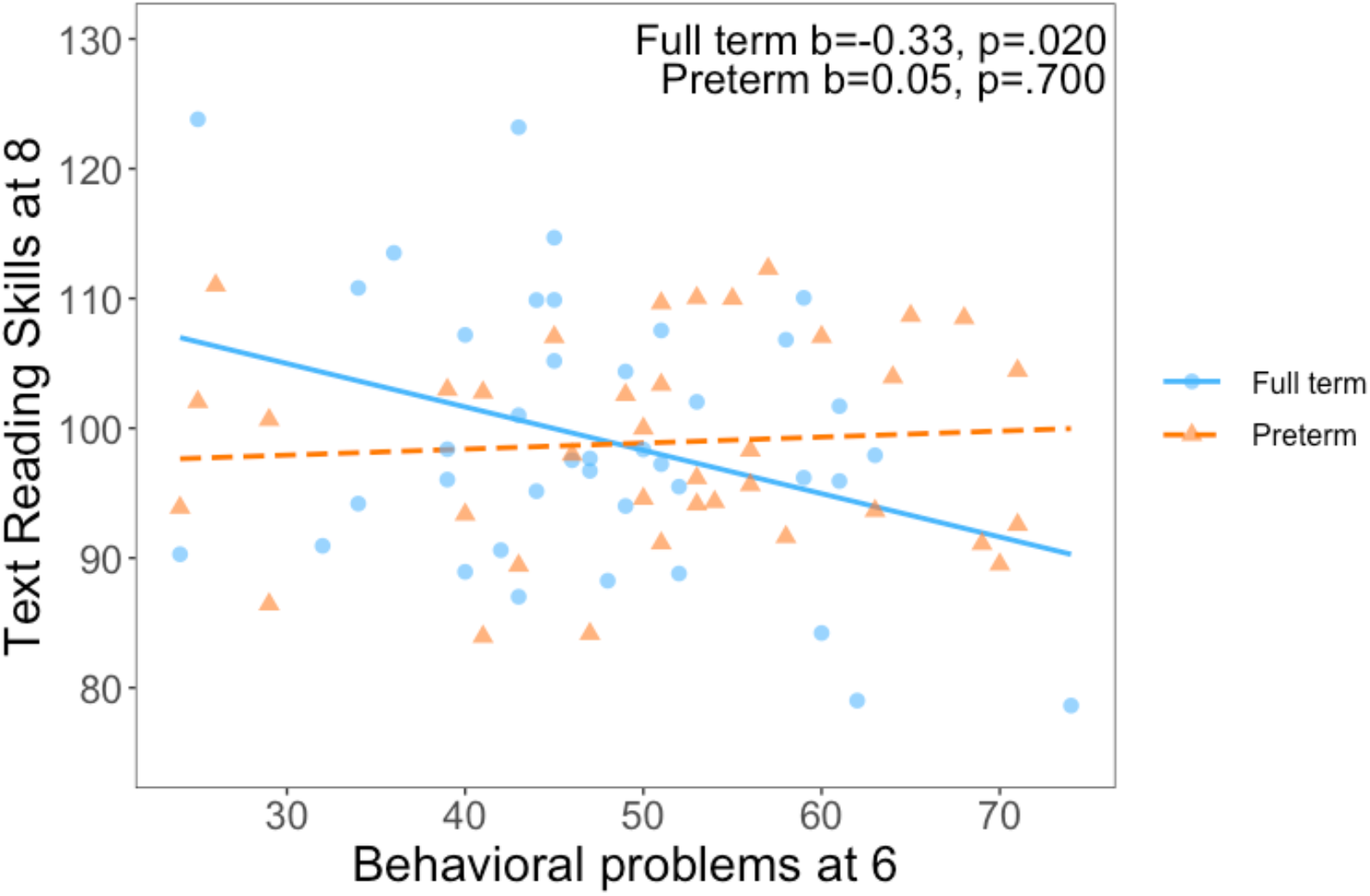
Regression residuals of behavioral problems at age 6 as a predictor of text reading skills at age 8 in children born at term and preterm, when accounting for sex and early reading skills at age 6.

## Discussion

The purpose of this study was to evaluate the longitudinal contribution of the severity of behavioral problems at age 6 to text reading skills at age 8, and if birth group moderated the association. We hypothesized we would find a negative association in the preterm group but not the term group. Our results demonstrated findings contrary to our hypothesis. In fact, we found that behavioral problems at age 6 were associated with text reading skills at age 8 only in the term group, not in the preterm group. This negative association persisted even when early reading skills at age 6 were considered in the model.

In the previous study of this cohort, we found that executive function and non-verbal IQ at age 6 improved the prediction of text reading skills at age 8 in children born preterm but not children born term[1]. We inferred that children born preterm relied on a broader set of early skills to read text than did children born term. However, in this analysis, we found that the association of behavioral problems and late text reading skills was present in the term group and not the preterm group.

Our results in the term group are similar to previous studies, which demonstrated that early externalizing problems are associated with late poor reading skills[8–10]. However, in our study, this association was between broader behavioral problems, and this association was not present in the preterm group. The results demonstrate the complexity of longitudinal prediction across clinical populations. One explanation may be that the nature of behavioral problems differs by birth group. Preterm children are more likely to experience internalizing problems rather than externalizing problems[11] as part of the preterm phenotype. Contrary to externalizing problems, association between early internalizing problems and late poor reading skills have been investigated in the literature, but the associations are rather mixed[9,10,12].

This study has several limitations. The sample size is limited and participants came primarily from high socioeconomic backgrounds[1]. Thus, the sample was not representative of many groups of children born preterm. Further, participants were recruited by their birth group status, not by behaviors, so the proportion of participants with severe behavioral problems was small. Behavioral problems were assessed using a parent-report measure only.

In conclusion, earlier domains of function outside of reading skills contribute to later reading skills in school-aged children, and the contributing domains vary by birth group; Borchers et al[1] found that earlier executive functioning and non-verbal intelligence contribute to later reading skills in children born preterm. Here we add that earlier total behavioral problems contribute to later reading skills only in children born term. Our findings have clinical implication that children born term with early behavioral problems need close monitoring of reading performance, regardless of their earlier reading skills competence. Future studies should seek to replicate our findings in a larger sample, ideally across multiple sites, and to broaden the scope by evaluating white matter properties as potential neurobiological mediators of the effects.

## Data Availability

All data produced in the present study are available upon reasonable request to the authors

## Funding Sources

This work was supported by National Institutes of Health (NICHD grant RO1-HD069162 A and 2RO1-HD069150) and Health Resources and Services Administration Maternal Child Health Bureau (T77MC09796) to Heidi M Feldman. Machiko Hosoki is the Charles B. Woodruff Endowed Fellow Of the Maternal Child Health Research Institute, Stanford University.

## Conflict of Interest Statement

None declared

## Notes

### Competing Interest Statement

The authors have declared no competing interest.

### Author Declarations

The Stanford University IRB gave ethical approval for this work

## References

[1] L.R. Borchers, L. Bruckert, K.E. Travis, C.K. Dodson, I.M. Loe, V.A. Marchman, H.M. Feldman, Predicting text reading skills at age 8 years in children born preterm and at term, Early Human Development. 130 (2019) 80–86.

[2] T.L. Blankenship, M.A. Slough, S.D. Calkins, K. Deater-Deckard, J. Kim-Spoon, M.A. Bell, Attention and executive functioning in infancy: Links to childhood executive function and reading achievement, Developmental Science. 22 (2019) e12824. https://doi.org/10.1111/desc.12824.

[3] C.S.H. Aarnoudse-Moens, N. Weisglas-Kuperus, J.B. van Goudoever, J. Oosterlaan, Meta-Analysis of Neurobehavioral Outcomes in Very Preterm and/or Very Low Birth Weight Children, Pediatrics. 124 (2009) 717–728. https://doi.org/10.1542/peds.2008-2816.

[4] T.M. Achenbach, C.S. Edelbrock, Manual for the Child: Behavior Checklist and Revised Child Behavior Profile, in: 1983.

[5] B. Larroque, S. Marret, P.-Y. Ancel, C. Arnaud, L. Marpeau, K. Supernant, V. Pierrat, J.-C. Rozé, J. Matis, G. Cambonie, A. Burguet, M. Andre, M. Kaminski, G. Bréart, White matter damage and intraventricular hemorrhage in very preterm infants: the EPIPAGE study, The Journal of Pediatrics. 143 (2003) 477–483. https://doi.org/10.1067/S0022-3476(03)00417-7.

[6] A.H. Hall, R.P. Tannebaum, Test Review: J. L. Wiederholt & B. R. Bryant. (2012). Gray Oral Reading Tests—Fifth Edition (GORT-5). Austin, TX: Pro-Ed., Journal of Psychoeducational Assessment. 31 (2013) 516–520. https://doi.org/10.1177/0734282912468578.

[7] D. Wechsler, WASI-II: Wechsler abbreviated scale of intelligence, PsychCorp, 2011.

[8] R.-L. Metsäpelto, G. Silinskas, N. Kiuru, A.-M. Poikkeus, E. Pakarinen, K. Vasalampi, M.-K. Lerkkanen, J.-E. Nurmi, Externalizing behavior problems and interest in reading as predictors of later reading skills and educational aspirations, Contemporary Educational Psychology. 49 (2017) 324–336. https://doi.org/10.1016/j.cedpsych.2017.03.009.

[9] J.V. der Ende, F.C. Verhulst, H. Tiemeier, The bidirectional pathways between internalizing and externalizing problems and academic performance from 6 to 18 years, Development and Psychopathology. 28 (2016) 855–867. https://doi.org/10.1017/S0954579416000353.

[10] S. Hagan-Burke, O. Kwok, Y. Zou, C. Johnson, D. Simmons, M.D. Coyne, An Examination of Problem Behaviors and Reading Outcomes in Kindergarten Students, J Spec Educ. 45 (2011) 131–148. https://doi.org/10.1177/0022466909359425.

[11] S. Johnson, N. Marlow, Preterm Birth and Childhood Psychiatric Disorders, Pediatric Research. 69 (2011) 11–18. https://doi.org/10.1203/PDR.0b013e318212faa0.

[12] B.P. Ackerman, C.E. Izard, R. Kobak, E.D. Brown, C. Smith, Relation Between Reading Problems and Internalizing Behavior in School for Preadolescent Children From Economically Disadvantaged Families, Child Development. 78 (2007) 581–596. https://doi.org/10.1111/j.1467-8624.2007.01015.x.

